# Hydroxychloroquine (HCQ): an observational cohort study in primary and secondary prevention of pneumonia in an at-risk population

**DOI:** 10.1101/2020.04.08.20057893

**Authors:** Alain Vanasse, Josiane Courteau, Yohann Chiu, André Cantin, Richard Leduc

## Abstract

**Background:** Recent studies suggest that hydroxychloroquine (HCQ) could be effective against COVID-19. It is reasonable to expect that if HCQ can prevent or reduce the adverse effects of influenza, it may also reduce the effects of COVID-19 in humans. The objective of this study was to test whether HCQ can prevent or reduce the risk and severity of influenza.

**Methods:** This is an observational cohort study using medico-administrative data from Québec. Patients included had at least one emergency department (ED) visit in 2012 or 2013, with a prior diagnosis of chronic conditions, and were admissible to the public drug insurance plan. Two sub-cohorts were considered depending on reasons for ED visit: other than influenza or pneumonia (primary prevention) and influenza or pneumonia (secondary prevention).

**Results:** In the primary prevention analysis (n=417,353), patients taking HCQ (n=3,659) had an increased risk of hospitalization for pneumonia in the following year compared to those who did not (5.2% vs. 2.9%; adjusted OR=1.25, p=0.0079). In the secondary prevention analysis (n=27,152), patients taking HCQ (n=392), compared to those who did not had a modest and non-significant increased risk of hospitalization for pneumonia after 30 days (25.8% vs. 22.6%; adjusted OR=1.14, p=0.3177).

**Interpretation:** Based on the assumption that HCQ has similar effects on the COVID-19 as those observed on influenza, we can infer that it will not have positive effects on COVID-19. We should therefore act cautiously before initiating prospective interventional studies on the use of HCQ to reduce adverse effects of COVID-19.

## INTRODUCTION

At this time, there is no treatment nor vaccine available to treat or prevent infection or symptoms associated with SARS-CoV-2. In this race against time, several treatment known to be effective on other viruses are currently being tested around the world in new randomized control trials.^1,2,3^ Meanwhile, recent *in vitro* studies suggest that hydroxychloroquine (HCQ) could be effective against coronavirus (COVID-19).^4^ The entry of the SARS-CoV-2 and influenza virus into host cells requires cleavage of the spike glycoprotein (SARS-CoV-2) and hemagglutinin-A (influenza) by host cell serine proteases.^5,6^ Host cell proprotein cleavage and host cell infection by influenza virus are both inhibited in vitro by endosomal alkalinizing agents.^7,8^ Based on these considerations, we put forward the hypothesis that if HCQ can prevent or reduce the adverse effects of influenza, it could prevent or reduce the harmful effects of COVID-19 in humans.

The objective of the present study is to test whether HCQ can prevent or reduce the risk and severity of infection by influenza. Respiratory hospitalization, mortality and influenza-like illness data as well as invasive pneumonia correlate with seasonal influenza and can be used as surrogate markers of influenza severity.^9^ More specifically, we address the following research questions: 1) In the at-risk population (primary prevention), do HCQ users have a lower rate of hospitalization for pneumonia compared to non-users? 2) Among at-risk individuals who visit the ED with a diagnosis of influenza or pneumonia (secondary prevention), do HCQ users have fewer hospitalizations for pneumonia in the following 30 days compared to non-users?

## METHODS

### Design and Data source

This is an observational retrospective cohort study using medico-administrative data from Québec (Canada). Medical services and drug reimbursement files, as well as hospitalization data, are held and managed by the *Régie de l’assurance maladie du Québec* (RAMQ), which provides universal health insurance to Quebec residents, including coverage for physician and hospital services. Using a unique encrypted identifier, patient data from these registers were linked to provide information on demographic characteristics, medical and prescription drug information.

### Study Cohort

Adult patients included in the study cohort had at least one emergency department (ED) visit (index date) between January 2012 and December 2013, with a prior diagnosis of ambulatory care sensitive condition (ACSC)^10^ and continuously admissible to the public prescription drug insurance plan (PPDIP) three months before the index date. Two sub-cohorts were considered separately depending on the reason for ED visit: other than influenza/pneumonia (primary prevention, ICD-9: other than 480-487, n= 417 353) and influenza/pneumonia (secondary prevention, ICD-9: 480-487, n=27 152).

### Variables

The binary dependent variable considered is the presence/absence of a hospitalization for pneumonia (ICD-10: J12-J18, J10.0, J11.0) within 365 days (primary prevention) or 30 days (secondary prevention) after index ED visit. For both analyses, the independent variable is at least one claim of HCQ within 3 months prior to index date (ED visit). The adjustment variables are age, sex, type of ACSC, a comorbidity index,^11^ and immunosuppressive medications for rheumatoid arthritis (prednisolone or prednisone, methotrexate, sulfasalazine, azathioprine, and cyclosporine).

### Statistical analysis

We used multiple logistic regression models to estimate adjusted odds ratios (OR). We performed several sensitivity analyses by modifying the study cohort (specific ICD code 487 for influenza as the reason of index ED visit) or the dependent variable (enlarging ICD coding for influenza/pneumonia and by including all-cause death).

### Ethics approval

This study was approved by the Research Ethics Board Committee of the Université de Sherbrooke and by the *Commission d’accès à l’information* of Quebec.

## RESULTS

The characteristics of the two cohorts are summarized in Table 1. Patients in the two cohorts are generally older that the general population (mean age 70 and 75 years) and represent a population with comorbidities (with at least one ACSC). An important proportion of patients in the secondary prevention cohort suffers from COPD.

**Table 1.** Characteristics of the study cohorts

|  | Primary prevention cohort |  | Secondary prevention cohort |  |
| --- | --- | --- | --- | --- |
| Characteristic | Total<br>(n=417 353) | Hospitalized for<br>pneumonia<br>n=12 194 (2.9%) | Total<br>(n=27 152) | Hospitalized for<br>pneumonia<br>n=6160 (22.7%) |
| Age, mean (SD) | 70.5 (14.3) | 77.3 (12.0) | 75.1 (13.2) | 77.9 (12.1) |
| Male sex, n (%) | 190 956 (45.8) | 5889 (48.3) | 13 103 (48.3) | 2981 (48.4) |
| ACSC - HBP, n (%) | 236 173 (56.6) | 7334 (60.1) | 16 873 (62.1) | 4071 (66.1) |
| ACSC - Diabetes, n (%) | 143 055 (34.3) | 4468 (36.6) | 9848 (36.3) | 2348 (38.1) |
| ACSC - CHD, n (%) | 118 449 (28.4) | 4750 (39.0) | 11 208 (41.3) | 2680 (43.5) |
| ACSC - COPD, n (%) | 70 348 (16.9) | 4568 (37.5) | 11 660 (42.9) | 2363 (38.4) |
| ACSC - CHF, n (%) | 34 629 (8.3) | 2356 (19.3) | 5829 (21.5) | 1461 (23.7) |
| ACSC - Asthma, n (%) | 33 839 (8.1) | 1117 (9.2) | 3322 (12.2) | 634 (10.3) |
| ACSC - Epilepsy, n (%) | 10 486 (2.5) | 326 (2.7) | 859 (3.2) | 201 (3.3) |
| Comorbidity index, n (%) |  |  |  |  |
| 0 | 215 104 (51.5) | 3604 (29.6) | 7982 (29.4) | 1617 (26.2) |
| 1-2 | 100 806 (24.2) | 3220 (26.4) | 6386 (23.5) | 1470 (23.9) |
| 3-4 | 42 538 (10.2) | 2037 (16.7) | 4525 (16.7) | 1066 (17.3) |
| ≥ 5 | 58 905 (14.1) | 3333 (27.3) | 8259 (30.4) | 2007 (32.6) |
| HCQ use, n (%) | 3659 (0.9) | 192 (2.9) | 392 (1.4) | 101 (1.6) |

In the primary prevention analysis (n=417 353), the 3,659 (0.9%) patients who were using HCQ at the ED visit were at higher risk of being hospitalized for pneumonia in the following year when compared to non-users (crude OR=1.85, p<0.0001; adjusted OR=1.25, p=0.0169) (Table 1). In the secondary prevention analysis (n=27 152), the 392 (1.4%) patients taking HCQ for medical reasons and diagnosed with influenza or pneumonia at their index ED visit had a non-significant increased risk of hospitalizations for pneumonia in the following 30 days compared to non-users (crude OR=1.19, p=0.1431; adjusted OR=1.14, p=0.3177) (Table 2). Several sensitivity analyses were performed (Tables 3 to 5) and they almost all come to the same conclusions (except when all-cause death is included in the outcome).

**Table 2.** Association (OR) between HCQ use and hospitalization for pneumonia

| PRIMARY PREVENTION (n=417 353) |  |  |  |  |  |
| --- | --- | --- | --- | --- | --- |
|  | Hospitalization for pneumonia 365 days (n=12 194) |  |  |  |  |
|  | Yes | No | Crude OR<br>(95% CI) p-value | Adjusted OR<sup>i</sup><br>(95% CI) p-value | Adjusted OR<sup>ii</sup><br>(95% CI) p-value |
| HCQ | 192 (5.2%) | 3467 (94.8%) | 1.85 (1.60 – 2.15)*** | 1.62 (1.39 – 1.88)*** | 1.25 (1.06 – 1.47)* |
| Non-HCQ | 12 002 (2.9%) | 401 692 (97.1%) |  |  |  |
| SECONDARY PREVENTION (n=27 152) |  |  |  |  |  |
|  | Hospitalization for pneumonia 30 days (n=6160) |  |  |  |  |
|  | Yes | No | Crude OR<br>(95% CI) p-value | Adjusted OR<br>(95% CI) p-value | Adjusted OR<br>(95% CI) p-value |
| HCQ | 101 (25.8%) | 291 (74.2%) | 1.19 (0.94 – 1.49) | 1.23 (0.98 – 1.55) | 1.14 (0.88 – 1.46) |
| Non-HCQ | 6059 (22.6%) | 20 701 (77.4%) |  |  |  |

\*\*\* p<0.0001; \*\* p<0.001; \* p<0.05
<sup>i</sup> adjusted for sex, age and comorbidities
<sup>ii</sup> adjusted for sex, age, comorbidities and co-medications

**Table 3.** Sensitivity analyses: cohort with an ED visit for another reason than influenza or pneumonia (n=417 353). Association (OR) between HCQ use and the outcome. In total, 3659 patients were taking HCQ.

| Outcome (365 days) | Crude OR | OR adjusted for sex, age and comorbidities | OR adjusted for sex, age, comorbidities and co-medications |
| --- | --- | --- | --- |
| Hosp. pneumonia (J12-J18, J10.0, J11.0) (n=12 194) | 1.85 (1.60 – 2.15)*** | 1.62 (1.39 – 1.88)*** | 1.25 (1.06 – 1.47)* |
| Among survivors | 1.90 (1.60 – 2.25)*** | 1.64 (1.38 – 1.94)*** | 1.26 (1.04 – 1.52)* |
| Hosp. influenza or viral pneumonia (J10-J12, J18) (n=11 734) | 1.78 (1.53 – 2.07)*** | 1.55 (1.33 – 1.81)*** | 1.22 (1.03 – 1.44)* |
| Among survivors | 1.80 (1.51 – 2.15)*** | 1.55 (1.30 – 1.85)*** | 1.22 (1.00 – 1.48)* |
| Hosp. influenza or pneumonia (J10-J18) (n=12 945) | 1.88 (1.63 – 2.16)*** | 1.64 (1.42 – 1.90)*** | 1.27 (1.08 – 1.48)* |
| Among survivors | 1.94 (1.64 – 2.28)*** | 1.67 (1.41 – 1.97)*** | 1.28 (1.07 – 1.54)* |
| Hosp. influenza or pneumonia (J10-J18) or death (n=52 302) | 1.39 (1.27 – 1.52)*** | 1.12 (1.02 – 1.23)* | 1.02 (0.92 – 1.13) |
| All-cause death (n=42 649) | 1.24 (1.12 – 1.37)*** | 0.98 (0.88 – 1.09) | 0.94 (0.84 – 1.06) |

\*\*\* p<0.0001; \*\* p<0.001; \* p<0.05

**Table 4.** Sensitivity analyses: cohort with an ED visit for influenza or pneumonia (n=27 152). Association (OR) between HCQ use and the outcome. In total, 392 patients were taking HCQ.

| Outcome (30 days) | Crude OR | OR adjusted for sex, age and comorbidities | OR adjusted for sex, age, comorbidities and co-medications |
| --- | --- | --- | --- |
| Hosp. pneumonia (J12-J18, J10.0, j11.0) (n=6160) | 1.19 (0.94 – 1.49) | 1.23 (0.98 – 1.55) | 1.14 (0.88 – 1.46) |
| Hosp. influenza or viral pneumonia (J10-J12, J18) (n=5805) | 1.10 (0.87 – 1.39) | 1.14 (0.90 – 1.45) | 1.06 (0.82 – 1.38) |
| Hosp. influenza or pneumonia (J10-J18) (n=6436) | 1.18 (0.94 – 1.48) | 1.23 (0.98 – 1.54) | 1.12 (0.88 – 1.44) |
| Hosp. influenza or pneumonia (J10-J18) or death (n=8364) | 1.17 (0.95 – 1.44) | 1.19 (0.96 – 1.47) | 1.13 (0.90 – 1.43) |

\*\*\* p<0.0001; \*\* p<0.001; \* p<0.05

**Table 5.**
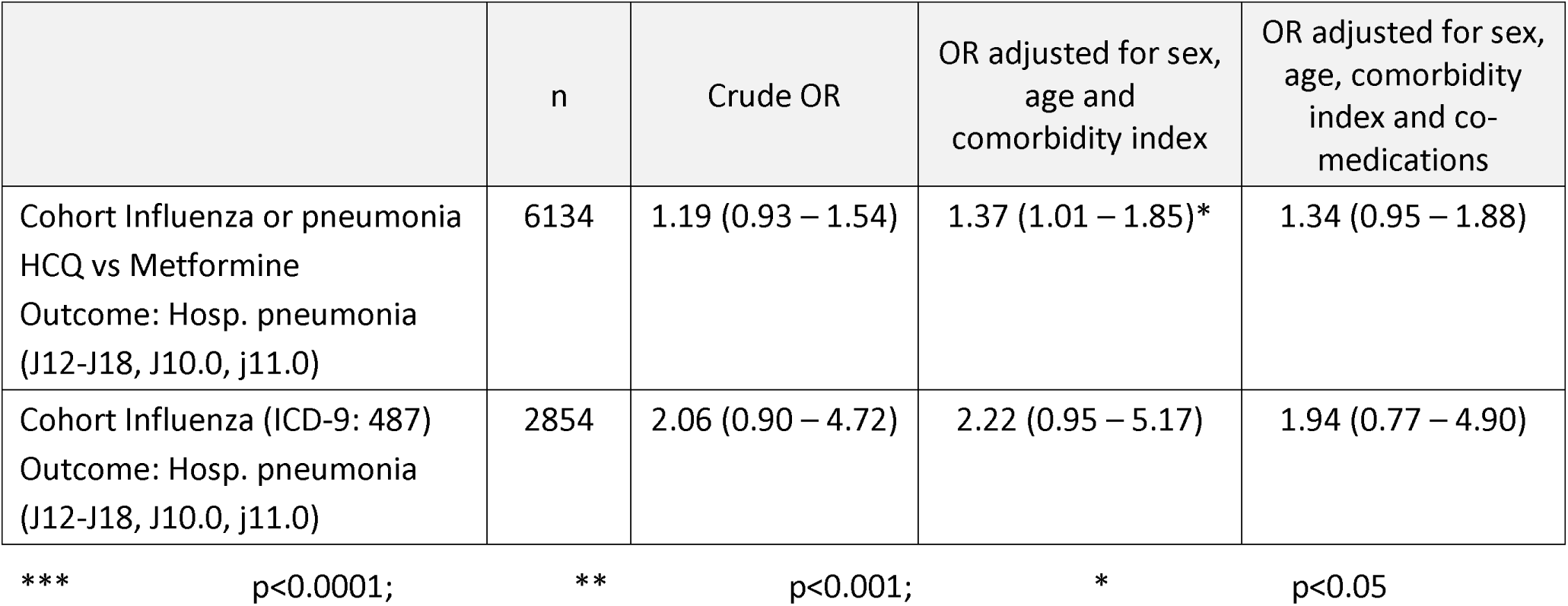
Additional sensitivity analyses for the secondary prevention cohort

|  | n | Crude OR | OR adjusted for sex, age and comorbidity index | OR adjusted for sex, age, comorbidity index and co-medications |
| --- | --- | --- | --- | --- |
| Cohort Influenza or pneumonia<br>HCQ vs Metformine<br>Outcome: Hosp. pneumonia (J12-J18, J10.0, j11.0) | 6134 | 1.19 (0.93 – 1.54) | 1.37 (1.01 – 1.85)* | 1.34 (0.95 – 1.88) |
| Cohort Influenza (ICD-9: 487)<br>Outcome: Hosp. pneumonia (J12-J18, J10.0, j11.0) | 2854 | 2.06 (0.90 – 4.72) | 2.22 (0.95 – 5.17) | 1.94 (0.77 – 4.90) |

\*\*\* p<0.0001; \*\* p<0.001; \* p<0.05

## INTERPRETATION

In primary prevention, there was an increased risk of 1-year hospitalization for pneumonia among HCQ users when compared to non-users, even after adjusting for age, sex, comorbidities and immunosuppressive co-medication (adjusted OR=1.25, p=0.0169) (Table 2). In secondary prevention, there was a modest and non-statistically significant increase of about 15% in the rate of hospitalization for pneumonia after 30 days of an ED visit for influenza or pneumonia (adjusted OR=1.14, p=0.3177) (Table 2). Multiple sensitivity analyses were performed, and the results pointed generally in the same direction (Tables 3-5). At first glance, HCQ appears to have adverse effects in primary and secondary prevention of influenza, but these adverse effects are reduced when adjusted analyses are performed and are probably due to the immunosuppressive co-medication received by patients taking HCQ for osteoarticular inflammatory problems.

HCQ has been shown to possess the ability to slow down progress of rheumatoid arthritis^12^ and good antiviral properties.^13^ In particular, chloroquine, of which HCQ is an analog, has been found effective against avian influenza in an animal model.^14^ However, the potential of HCQ and chloroquine in influenza prevention in humans is unclear at the moment.^15,16,17^ In some cases, those drugs may even be detrimental. For instance, the American College of Rheumatology recommends influenza vaccination for patients with rheumatoid arthritis before HCQ treatment.^18^

### Strengths and Limitations

The study has good external validity for Quebec since it is an exhaustive sample in a real-life context, however the results are only transferable to the population with ACSC and consulting EDs. The sensitivity of the study sample may be sub-optimal in that patients do not always consult a physician at ED for influenza or pneumonia or if they do, they may receive an inaccurate diagnosis and are not included in this study. On the other hand, specificity should be good, as the codes used are relatively specific for influenza or pneumonia, meaning that patients identified as having influenza in the study were likely to have had influenza. Although the drug codes are valid and reliable, Quebec’s PPDIP only covers a portion of the population (around 45%), namely patients aged 65 and over and those not covered by a private drug insurance plan; in short, an older and disadvantaged population. Prescribing bias is possible when comparing drug users with non-users. However, since there is no recommendation for influenza-pneumonia treatment that includes HCQ, it is unlikely that such bias systematically influenced the study results. Diagnostic codes on RAMQ reimbursement forms have been validated several times, but not for influenza, to our knowledge. The immortality bias is always possible in observational studies, but additional analyses (removing patients that died during the follow-up period) have been performed and do not suggest the presence of this bias. A confounding bias may also be present which could explain part of the results by an influence external to the study and affecting the study group without influencing the control group or vice versa. HCQ is often used in combination with other medications to reduce inflammation in patients with osteoarticular disease.^19,20^ Many of these drugs make patients more susceptible to viral infections including influenza and its associated conditions. Analyses adjusted for this type of bias were performed to account for it in the results.

## Conclusion

In conclusion, as no significant correlation between HCQ and benefits in primary or secondary prevention of influenza was observed in our study, researchers should be cautious when considering interventional studies on HCQ and COVID-19 with frail elderly people, a population particularly affected by COVID-19 but also more prone to experience adverse effects of HCQ.^21,22^

## Data Availability

The datasets generated and/or analysed during the current study are not publicly available due to individual privacy but are available from the corresponding author on reasonable request.

## Acknowledgements

This project was made possible thanks to the availability of a medico-administrative database managed by the Alain Vanasse and Catherine Hudon team (co-investigators of a Fonds de recherche du Québec—Santé grant).

## Footnotes

i adjusted for sex, age and comorbidities

ii adjusted for sex, age, comorbidities and co-medications

